# Viral infection and transmission in a large, well-traced outbreak caused by the SARS-CoV-2 Delta variant

**DOI:** 10.1101/2021.07.07.21260122

**Authors:** Baisheng Li, Aiping Deng, Kuibiao Li, Yao Hu, Zhencui Li, Qianling Xiong, Zhe Liu, Qianfang Guo, Lirong Zou, Huan Zhang, Meng Zhang, Fangzhu Ouyang, Juan Su, Wenzhe Su, Jing Xu, Huifang Lin, Jing Sun, Jinju Peng, Huiming Jiang, Pingping Zhou, Ting Hu, Min Luo, Yingtao Zhang, Huanying Zheng, Jianpeng Xiao, Tao Liu, Rongfei Che, Hanri Zeng, Zhonghua Zheng, Yushi Huang, Jianxiang Yu, Lina Yi, Jie Wu, Jingdiao Chen, Haojie Zhong, Xiaoling Deng, Min Kang, Oliver G. Pybus, Matthew Hall, Katrina A. Lythgoe, Yan Li, Jun Yuan, Jianfeng He, Jing Lu

## Abstract

We report the first local transmission of the SARS-CoV-2 Delta variant in mainland China. All 167 infections could be traced back to the first index case. Daily sequential PCR testing of the quarantined subjects indicated that the viral loads of Delta infections, when they first become PCR+, were on average ∼1000 times greater compared to A/B lineage infections during initial epidemic wave in China in early 2020, suggesting potentially faster viral replication and greater infectiousness of Delta during early infection. We performed high-quality sequencing on samples from 126 individuals. Reliable epidemiological data meant that, for 111 transmission events, the donor and recipient cases were known. The estimated transmission bottleneck size was 1-3 virions with most minor intra-host single nucleotide variants (iSNVs) failing to transmit to the recipients. However, transmission heterogeneity of SARS-CoV-2 was also observed. The transmission of minor iSNVs resulted in at least 4 of the 30 substitutions identified in the outbreak, highlighting the contribution of intra-host variants to population level viral diversity during rapid spread. Disease control activities, such as the frequency of population testing, quarantine during pre-symptomatic infection, and level of virus genomic surveillance should be adjusted in order to account for the increasing prevalence of the Delta variant worldwide.

---

During the global spread of the COVID-19, genetic variants of the SARS-CoV-2 virus have emerged. Some variants have increased transmissibility or could exhibit an increased propensity for escape from host immunity, and therefore pose an increased risk to global public health^1–3^. An emerging genetic lineage, B.1.617, has gained global attention and has been dominant in the largest outbreak of COVID-19 in India since March 2021. One descendent lineage, B.1.617.2, which carries spike protein mutations L452R, T478K and P681R, accounts for ∼28% sequenced cases in India and has rapidly replaced other lineages to become dominant in multiple regions and countries (https://outbreak.info/)^4^. Lineage B.1.617.2 has been labeled a variant of concern (VOC) and given the name Delta (https://www.who.int/activities/tracking-SARS-CoV-2-variants). Data on the virological profile of the Delta VOC is urgently needed.

On May 21, 2021 the first local infection of the Delta variant in Guangzhou, Guangdong, China was identified. As of the early epidemic in China in January 2020^5^, a suite of comprehensive interventions have been implemented to limit transmission, including population screening, active contact tracing, and centralized quarantine/isolation. However, in contrast to the limited level of onward transmission observed in Guangdong in early 2020^5^, successive generations of virus transmission were observed in the 2021 outbreak of the Delta variant in the region. Here, we investigated epidemiological and genetic data from the well-traced outbreak in Guangdong in order to characterize the virological and transmission profiles of the Delta variant. We discuss how intervention strategies may need to be adjusted to cope with the virological properties of this emerging variant.

## Results

A total of 167 local infections were identified during the outbreak, starting with the first index case identified on May 21, 2021 and ending with the last case reported on June 18, 2021 (Figure 1a). All cases could be epidemiologically or genetically traced back to the first index case (Figure 1b). One notable epidemiologic feature of the Delta variant is a shorter serial interval compared with to infection with early Wuhan-like strains or other VOC variants^6–8^. However, critical parameters before the illness onset remain poorly known, including when the viruses can be first detected in a subject after exposure, and how infectious infected individuals are.

**Figure 1:**
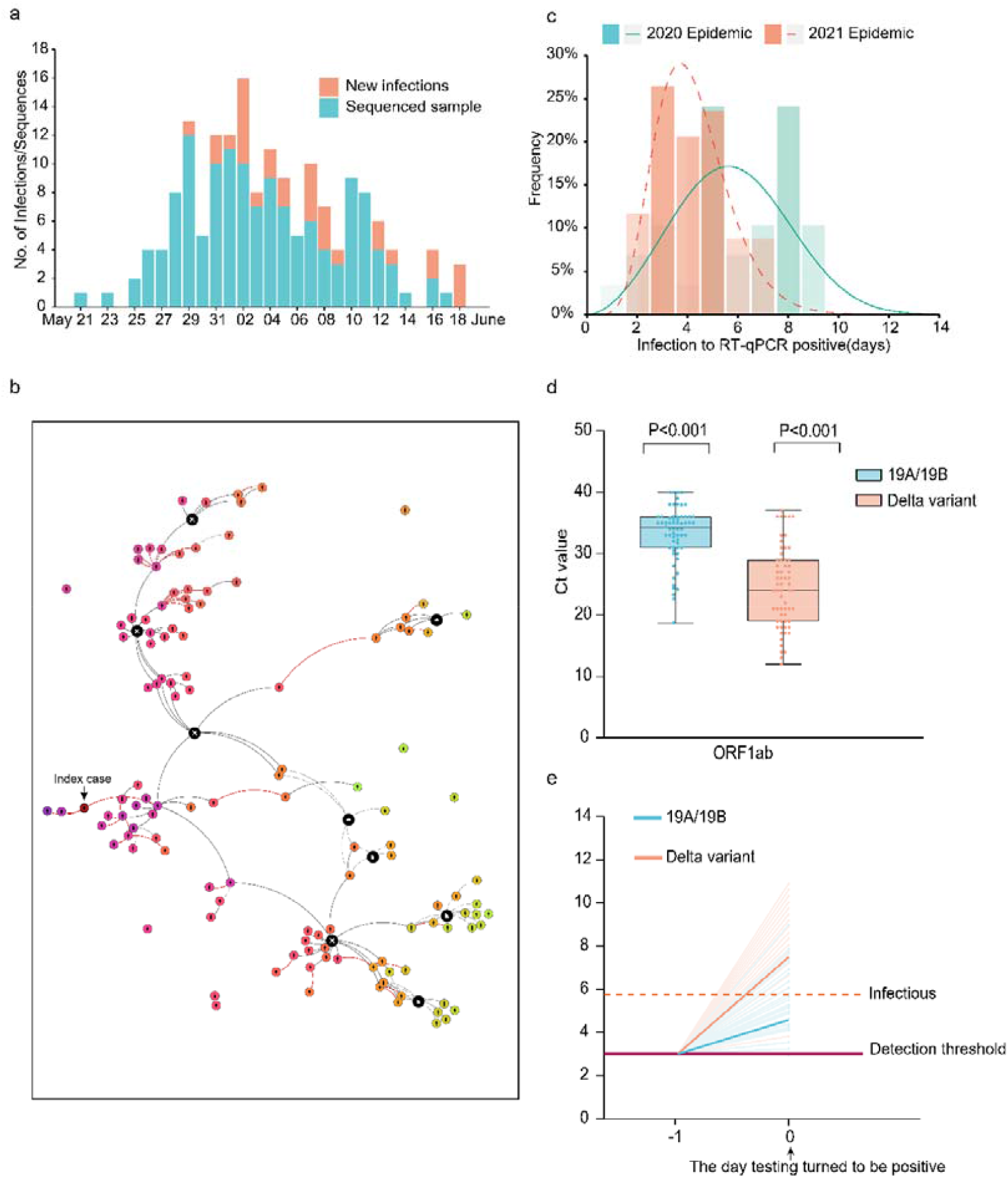
Summary of the epidemiology and early detection of the Delta SARS-CoV-2 variant in Guangdong. (a) Time series of 167 laboratory-confirmed infections originating from the first index case on May 21 2021. Daily numbers of new infections are shown in red and samples with high-quality sequences (coverage>95%) are shown in blue. (b) The Delta variant transmission in the Guangzhou outbreak. The transmission relationship between 126 sequenced cases were indicated with solid lines (high confidence) or the dash lines (unsure). The interactive version showing summary statistics of all cases could be found at https://viz.vslashr.com/guangdongcdc/. (c) Estimate of the time interval between exposure and time of the first RT-PCR positive test in quarantined subjects. The curves show the best-fitting distributions of the interval durations for Delta variant cases (n=34) and for 19A/19B clade cases (n=29). Bars show the histograms of estiamted intervals durations (days). (d) Ct values of the first PCR+ test in quarantined subjects, for the Delta variant infections (n=62) and for previous 19A/19B clade strains infections (n=63). Dots represent Ct values for RT-PCR of the ORF1ab gene (left) and N gene (right). Box plots indicate the median and interquartile range (IQR); the whiskers represent the maximum and minimum values. Schematic of the relation between the viral growth rate and the relative viral loads on the day viruses were first detected (Day 0). The viral load of A/B clade infections and of the Delta variant infections on Day 0 were measured. The horizontal dashed line in purple represents the detection threshold of RT-PCR testing; the dashed line in red represents the lower limit above which infectious viruses could be potentially isolated.

We investigated the data from the quarantined subjects in this outbreak and compared it to data from the early 2020 epidemic caused by A/B genetic clade (Pango nomenclature^9^) strains. The centrally-quarantined subjects were the close contacts of confirmed cases. Once a new infection was identified, his/her close contacts were immediately traced, centrally isolated, and underwent daily PCR testing. The dataset from quarantined subjects allowed us to determine the time interval in the infected subjects between exposure and when viral loads were first detectable by PCR. The exact exposure time for the intra-family transmissions was difficult to pinpoint, hence we removed intra-family transmission pairs from our time interval analysis. Our results revealed that the time interval from exposure to the first PCR+ test in the quarantined population was 6.00 days (IQR 5.00-8.00) during the 2020 epidemic (n=29; peak at 5.61 days) and 4.00 days (IQR 3.00-5.00) in the 2021 Delta epidemic (n=34; peak at 3.71 days; Figure 1c).

We next evaluated viral load measurements at the time when SARS-CoV-2 was first detected by PCR in each subject. The relative viral loads of cases infected with the Delta variant (n=62, Ct =24.00 for the *ORF1ab* gene, IQR 19.00∼29.00) were 1260 times higher than those for the 2020 infections with clade 19A/19B viruses (n=63, Ct = 34.31 for *ORF1ab* gene, IQR 31.00∼36.00) on the day when viruses were first detected (Figure 1d). We hypothesized a higher within-host growth rate of the Delta variant, which led to the higher observed viral loads once viral nucleotides exceeded the PCR detection threshold (Figure 1e). Similar to results reported by Roman *et*.*al*., we found that samples with Ct > 30 (<6×10^5^ copies/mL viruses) did not yield an infectious isolate in-vitro. For the Delta variant infections, 80.65% of samples contained >6×10^5^ copies/mL in oropharyngeal swabs when the viruses were first detected, compared to 19.05% of samples from clade 19A/19B infections. These data indicate that the Delta variant could be more infectious during the early stage of the infection (Figure 1e).

Individuals undergo a latent period after infection, during which viral titers are too low to be detected. As viral proliferation continues within host, the viral load will eventually reach detectable levels and the individual will become infectious. Knowing when an infected person can transmit is essential for designing intervention strategies that break chains of transmission. However, infectiousness is difficult to measure from clinical investigations since >50% of transmission occurs during the pre-symptomatic phase^10^. Our investigation of quarantined subjects suggests that, for the Delta variant, the time window from exposure to the detection of virus was ∼3.7 days, and infections presented a higher transmission risk when the virus was first detected compared to earlier circulating viral lineages. Consequently, the provincial government required people leaving Guangzhou city from airports, train stations and shuttle bus stations to show proof of a negative COVID-19 test within 72 hours on June 6 and this was shortened to 48 hours on June 7. In contrast, the comparable time window implemented in the 2020 epidemic was seven days.

### Transmission bottleneck and the association between minor iSNVs transmissions and viral population diversity

The non-pharmaceutical interventions in Guangdong mainly focus on epidemiological investigation, contact tracing and mass testing. Approximately 30 million PCR tests were performed between May 26, 2021 and June 8, 2021. The intense testing and screening of high-risk populations makes cryptic transmissions unlikely. Nearly all the infections we identified could be connected epidemiologically, either through evidence of direct contact, or indirectly (staying in or visiting the same area) (Figure 1b). In addition, all sequences could be genetically traced back to the index case. This provided a unique opportunity for us to characterize virus transmission dynamics at a finer scale, particularly the extent to which virus genetic diversity is transmitted among hosts. Whole-genome deep sequencing was performed on all identified infections, and 126 high-quality viral genomes (coverage>95%) were obtained, comprising 75% of identified infections in the outbreak (Figure 1a).

Phylogenetic analysis was performed by combining the virus genomes we obtained from the Delta outbreak with genomes from 346 imported cases; the latter represent travelers to Guangdong during March 2020 to June 2021 who arrived from 66 different source countries. We also included a set of reference sequences, comprising 50 genomes randomly selected from each of 13 defined NextStrain clades (https://nextstrain.org/) and the notified VOCs (Alpha, Beta, Gamma, Delta). The viral lineage distribution of the imported cases was approximately representative of the SARS-CoV-2 genetic lineages that were circulating at that time at the global scale. These importations pose a challenge for disease control and prevention in Guangdong, China (Figure 2a).

**Figure 2:**
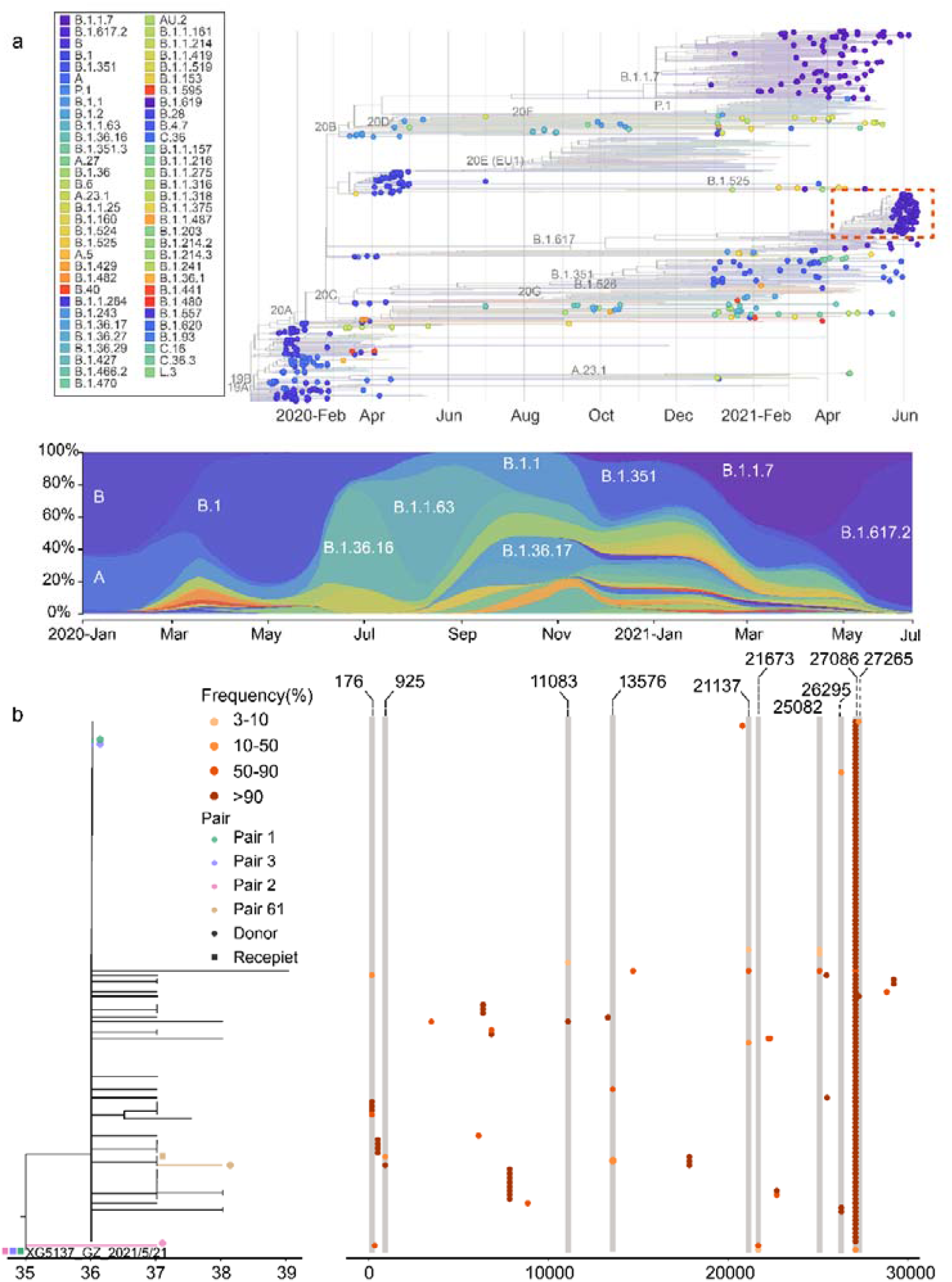
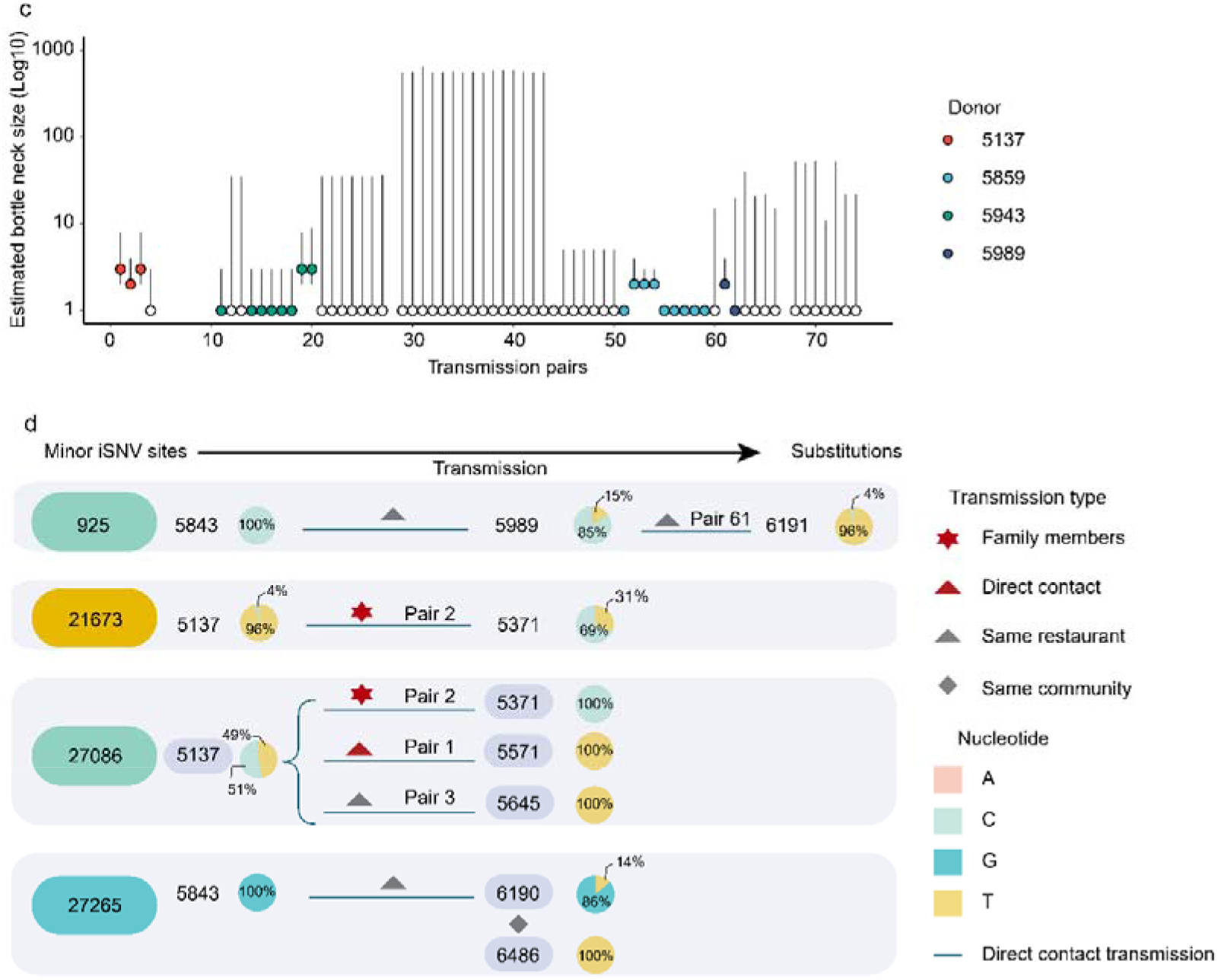
Viral phylogenies and transmission dynamics of the Guangzhou outbreak. (a) A time resolved phylogenetic tree was estimated using the NextStrain pipeline and includes (i) Guangdong sequences collected from local infections and imported cases, January 2020 – June 2021, and (ii) reference sequences from different genetic lineages. The sequences from the Guangdong Delta variant outbreak (May 21,2021 – June 18, 2021) are highlighted with a red box. The changing frequencies of SARS-CoV-2 lineages identified in Guangdong (most of which are imported) are shown in the lower panel (b) Maximum likelihood tree of 126 sampled sequences of the Guangzhou outbreak. SNV frequencies (%) across the virus genome are marked with colored dots (right hand panel). (c) Estimated bottleneck size in 66 donor-recipient transmission pairs calculated using the exact beta-binomial method described in ^14^. There were 8 transmission pairs with extremely large confidence interval (range from 1 to more than 1000) of estimated bottleneck size were removed. (d) Minor iSNVs transmission resulted in the diversity of viral population. The pie charts show the frequency of iSNVs. Arrows show the direction of transmission for those pairs of cases for which this is known with high confidence.

Viral phylogenies of the Guangzhou outbreak were inferred using the assembled consensus sequence of each sample, which was generated by choosing the majority-frequency nucleotide (>50%) at each position. All Guangzhou outbreak sequences segregated into a single cluster (Figure 2a). Compared with the index case (5137) of the outbreak, 30 substitutions were identified among 125 cases during the 26-days long outbreak (Figure 2b). The most genetically-divergent outbreak sequence contained four nucleotide differences from the index case sample. To understand how these variants emerged, grew and finally fixed during the epidemic (and during the SARS-CoV-2 pandemic more generally), we estimated within-host virus diversity for each sample by mapping polymorphic sites against the consensus genome of the index case (XG5137_GZ_2021/5/21), thereby generating a list of intra-host single-nucleotide variants (iSNVs). Minor iSNVs were called by setting 3% as the threshold for minor allele frequency, in order to exclude potential PCR and sequencing errors ^11–13^. For 126 high-quality sequences, most samples harbored 3 iSNVs (median) which is consistent with other reported levels (Supplemental Figure) ^11,12^.

We calculated the transmission bottleneck size among epidemiologically-confirmed transmission pairs. Contact tracing and epidemiological investigation enabled us assign 111 donor-recipient transmission pairs with a high degree of confidence. Of these, the donor had one or more iSNVs above the variant calling threshold of 3% in 74 transmission pairs (Table S1), enabling estimation of the transmission bottleneck size, *N*_*b*_, using the beta-binomial method ^14^. The maximum likelihood estimate for *N*_*b*_ was one for 65 out of these 74 transmission pairs, and two or three for the remaining 9 transmission pairs (Figure 2C). Uncertainty in the *N*_*b*_ estimate was large for some transmission pairs, with the 95% confidence interval ranging from 1 to ∼500 or more, suggesting for some pairs the sequencing data was not sufficiently informative. Our data suggest the transmission bottleneck of SARS-CoV-2 is very narrow in general, consistent with the previous household transmission studies ^11,15^. The transmission bottleneck size influences the extent to which within-host diversity contributes to viral diversity at the population scale. The stringent transmission bottleneck of SARS-CoV-2 suggests the substitutions we observed in Guangdong outbreak (and SARS-CoV-2 pandemic more generally) largely resulted from de-novo mutations appearing within individuals.

Although the transmission bottleneck of SARS-CoV-2 is narrow in general, it may be not constant and could be impacted by both viral and host factors. To investigate the contribution of the transmission of minor iSNV to population-level diversity, we identified the sequences with minor iSNVs and the sequences in which the derived nucleotide state was fixed. Notably, sequences exhibited minor intra-host single nucleotide variants (iSNVs) at 10 of the 30 variant sites (positions that varied from the sequence of the first index case) (Figure 2b). The direct (transmission pair 61, 1, 2, 3) and indirect (from case 6190 to 6486) epidemiological links were observed between the hosts with the minor iSNVs and their potential recipients with these iSNVs fixed (Figure 2C). Therefore, at least three fixed substitutions in this outbreak could be traced to the direct transmission of minor iSNVs, and one substitution was from a suspicious transmission chain. It is also noteworthy that the transmission pairs with 5137 as the donor had a relatively higher estimated *N*_*b*_, suggesting heterogenicity in iSNV transmission (Figure 2c). The differences in bottleneck size are possibly due to the different transmission route or exposure doses, as has been observed for influenza ^16^. The case 5137 presented a high viral load (Ct value of 17.6, approximate 2×10^9^ copies/mL in oropharyngeal swabs) 2 days after their direct contact with the cases 5645 and 5571. The high viral loads, direct contacts and relatively high frequency of the iSNVs (4% for T21673C and 47% for C27086T) may have enabled the successful transmission of iSNVs to the recipients (Figure 2C). Taken together, our observations suggest that the transmission bottleneck of SARS-CoV-2 is stringent in general, with most donor iSNVs not found in the recipients. However, transmission of minor iSNVs, with their fixation in the recipient host, resulted in at least some of the substitutions that accumulated during the outbreak.

In this study, we characterized a large transmission chain that originated from the first local infection of the SARS-CoV-2 Delta variant in mainland China. We find evidence for a potentially higher viral replication rate of the Delta variant, as viral loads in Delta infections are ∼1000 times higher than those for clade 19A/19B infections on the day of the first PCR+ test. This suggests that infectiousness of Delta variant during the early stage of infection is likely to be higher. Consequently, the frequency of population screening should be optimized ^17^. If Delta infections are indeed more infectious during the pre-symptomatic phase, then timely quarantine (before clinical onset or PCR screening) for suspected cases or for close contacts becomes more important. Although the transmission bottleneck of SARS-CoV-2 is narrow in general, heterogenicity of minor iSNV transmission is observed and explains some of the fixed substitutions observed in the virus population during the outbreak. In some settings, the advantageous iSNVs that are present at a low frequency could rise and become fixed in the one generation of transmission, and further predominance in the virus population if the epidemic is not well contained.

## Methods

### Ethics

This study was approved by the institutional ethics committee of the Guangdong Provincial Center for Disease Control and Prevention (GDCDC). Written consent was obtained from patients or their guardian(s) when samples were collected. Patients were informed about the surveillance before providing written consent, and data directly related to disease control were collected and anonymized for analysis.

### Sample collection, clinical surveillance and epidemiological data

Since the first local SARS-CoV-2 infection reported on May 21 in the capital city of Guangdong, the enhanced surveillance was performed by Guangdong CDC and local CDCs to detect suspected infections. Epidemiological investigations had been done on all confirmed cases. Population screening were performed by third-party detection institutions. Once virus positive samples were confirmed by local CDCs or other institutions, the samples were required to send to Guangdong CDC in 24 hours. To make the results comparable, in Guangdong CDC, the real-time reverse transcription

PCR (RT-PCR) were performed by using the same commercial kit (DaAn Gene) and RT-PCR machine (CFX96) as the previous studies^5,18^. The exposure history for positive cases and their close contacts were obtained through an interview, public video monitoring systems and cell phone apps, *etc*. Information regarding the demographic and geographic distribution of SARS-CoV-2 cases can be found at the website of Health Commission of Guangdong Province (http://wsjkw.gd.gov.cn/xxgzbdfk/yqtb/). The surge population screening test ensure all possible infections were identified and 111 donor-recipient transmission pairs were assigned with very high confidence. All transmission pairs met the following rules: 1. The recipient was the close contract of the donor and had a clear and direct epidemiological link to the donor; 2. The recipient did not have any contacts with other identified cases.

### Virus amplification and sequencing

Total RNAs were extracted from oropharyngeal swab samples by using QIAamp Viral RNA Mini Kit (Qiagen, Cat. No. 52904). Virus genomes were generated by two different approaches, (i) using commercial sequencing kit of BGI (ATOPlex 1000021625) and sequencing on the BGI MGISEQ-2000 (n=25), and (ii) using version 3 of the ARTIC COVID-19 multiplex PCR primers (https://artic.network/ncov-2019) for genome amplification, followed by library construction with Illumina Nextera XT DNA Library Preparation Kit and sequencing with PE150 (n=63) or SE100 (n=38) on Illumina Miniseq. We report only high-quality genome sequences for which we were able to generate >95% genome coverage.

### Sequence analysis

The bioinformatics pipeline for BGI platform (https://github.com/MGI-tech-bioinformatics/SARS-CoV-2_Multi-PCR_v1.0) was used to generate consensus sequences and call single nucleotide variants relative to the reference sequence. For sequence data from Miniseq, the raw data were first quality controlled (QC) using fastp^19^ to trim artificial sequences (adapters), to cut low-quality bases (quality scores <□20). PCR primers were trimmed by using cutadapt version 3.1^20^ or other published method^21^. Since all infections could be traced back to the first index case, the cleaned reads of each sample were mapped against the genome of the first index case (5137_GZ_2021/5/21) using BWA 0.7.17^22^. The consensus sequences were determined with iVar 1.2.1^23^, taking the most common base as the consensus (allele frequency >50%). An N was placed at positions along the reference with the sequencing depth fewer ≤ 10. The surge population screening test ensure all possible infections were identified and through the contact tracing the donor-recipient transmission pairs could be assigned with high confidence. To characterize the viral transmission in these pairs, we identified iSNVs relative to the reference genome (XG5137_GZ_2021/5/21) for each sequence with iVar 1.2.1 using the following parameters: alternated frequency at a SNV site ≥ 3%; total sequencing depth at SNV site ≥ 100; sequencing depth for the variant allele ≥ 10; iVar PASS=TRUE. We exclude the head and tail sequences of viral genome (corresponding to the positions 1 to 100 and 29803 to 29903 in Wuhan-Hu-1 reference genome) due to the lower sequencing coverage for most samples in the analysis and the 7 “highly shared” iSNV sites (1959, 4091, 21987, 24404, 28448, 28389, 29681) possibly due to the contamination of the primer sequences or mapping errors ^11^. To infer the iSNVs transmission in 74 donor-recipient pairs, all sites with ≥3% minor allele frequency in the assumed donor were used in the analysis. In the recipient, all reads at these sites were considered, with a variant calling threshold of 3% using the beta-binomial method of Sobel Leonard *et*.*al*^14^. The nextstrain pipeline^24^ was used to analyze and visualize the genetic distribution of SARS-CoV-2 infections and its dynamic change in Guangdong between January 2020 and June 2021. Maximum likelihood (ML) tree was estimated with phyml^25^ using the HKY+Q4 substitution model with gamma-distributed rate variation^26^. The branch length was recalculated as the number of mutations to the reference sequence of the first index case. The tree was visualized with R package of ggtree^27^.

## Data Availability

All sequencing reads after primer trimming and mapped to the reference sequence (the sequences of the first index case, XG5137_GZ_2021/5/21) have been submitted to the National Genomics Data Center (https://bigd.big.ac.cn/) with submission number CRA004571. The generated consensus sequences were submitted with accession number GWHBDIM01000000-GWHBDNH01000000.

## Data availability

All sequencing reads after primer trimming and mapped to the reference sequence (the sequences of the first index case, XG5137_GZ_2021/5/21) have been submitted to the National Genomics Data Center (https://bigd.big.ac.cn/) with submission number CRA004571. The generated consensus sequences were submitted with accession number GWHBDIM01000000 – GWHBDNH01000000.

## Code availability

The pipeline for sequencing data analysis was deposit in https://github.com/Jinglu1982/Delta-variant-outbreak-in-GZ. Code to implement the beta-binomial method is publicly available^14^.

## Acknowledgements

We gratefully acknowledge the efforts of China national CDCs, Guangdong local CDCs, hospitals, and the third-party detection institutions in epidemiological investigations, sample collection, and detection. This work was supported by grants from Science and Technology Planning Project of Guangdong (2018B020207006), the Key Research and Development Program of Guangdong Province (2019B111103001), and Guangdong Workstation for Emerging infectious Disease Control and Prevention, Chinese Academy of Medical Sciences (2020-PT330-004).

## Competing interests

The views expressed in this article are those of the authors and not necessarily those of the Guangdong Provincial Center for Diseases Control and Prevention, or the Guangdong Provincial Institute of Public Health.

## Notes

### Funding Statement

1.Science and Technology Planning Project of Guangdong (2018B020207006)
2.The Key Research and Development Program of Guangdong Province (2019B111103001)
3.Guangdong Workstation for Emerging infectious Disease Control and Prevention, Chinese Academy of Medical Sciences (2020-PT330-004)

### Author Declarations

This study was approved by the institutional ethics committee of the Guangdong Provincial Center for Disease Control and Prevention (GDCDC). Written consent was obtained from patients or their guardian(s) when samples were collected. Patients were informed about the surveillance before providing written consent, and data directly related to disease control were collected and anonymized for analysis. For immunity evaluation, serum samples were collected from vaccinated volunteers in GDCDC with written consent.

### Summary of Updates

We report the first local transmission of the SARS-CoV-2 Delta variant in mainland China. All 167 infections could be traced back to the first index case. Daily sequential PCR testing of the quarantined subjects indicated that the viral loads of Delta infections, when they first become PCR+, were on average ~1000 times greater compared to A/B lineage infections during initial epidemic wave in China in early 2020, suggesting potentially faster viral replication and greater infectiousness of Delta during early infection. We performed high-quality sequencing on samples from 126 individuals. Reliable epidemiological data meant that, for 111 transmission events, the donor and recipient cases were known. The estimated transmission bottleneck size was 1-3 virions with most minor intra-host single nucleotide variants (iSNVs) failing to transmit to the recipients. However, transmission heterogeneity of SARS-CoV-2 was also observed. The transmission of minor iSNVs resulted in at least 4 of the 30 substitutions identified in the outbreak, highlighting the contribution of intra-host variants to population level viral diversity during rapid spread. Disease control activities, such as the frequency of population testing, quarantine during pre-symptomatic infection, and level of virus genomic surveillance should be adjusted in order to account for the increasing prevalence of the Delta variant worldwide.

## References

1. Faria, N. R. et al. Genomics and epidemiology of the P.1 SARS-CoV-2 lineage in Manaus, Brazil. Science (2021) doi:10.1126/science.abh2644.

2. Davies, N. G. et al. Estimated transmissibility and impact of SARS-CoV-2 lineage B.1.1.7 in England. Science 372, (2021).

3. Volz, E. et al. Assessing transmissibility of SARS-CoV-2 lineage B.1.1.7 in England. Nature 593, 266–269 (2021).

4. outbreak.info.

5. Lu, J. et al. Genomic Epidemiology of SARS-CoV-2 in Guangdong Province, China. Cell S0092867420304864 (2020) doi:10.1016/j.cell.2020.04.023.

6. Vöhringer, H. S. et al. Genomic reconstruction of the SARS-CoV-2 epidemic across England from September 2020 to May 2021. http://medrxiv.org/lookup/doi/10.1101/2021.05.22.21257633 (2021) doi:10.1101/2021.05.22.21257633.

7. Pung, R., Mak, T. M., CMMID COVID-19 working group, Kucharski, A. J. & Lee, V. J. Serial intervals observed in SARS-CoV-2 B.1.617.2 variant cases. http://medrxiv.org/lookup/doi/10.1101/2021.06.04.21258205 (2021) doi:10.1101/2021.06.04.21258205.

8. Zhang, M. et al. Transmission Dynamics of an Outbreak of the COVID-19 Delta Variant B.1.617.2 — Guangdong Province, China, May–June 2021. China CDC Wkly. 3, 584–586 (2021).

9. Rambaut, A. et al. A dynamic nomenclature proposal for SARS-CoV-2 lineages to assist genomic epidemiology. Nat. Microbiol. 5, 1403–1407 (2020).

10. Hu, S. et al. Infectivity, susceptibility, and risk factors associated with SARS-CoV-2 transmission under intensive contact tracing in Hunan, China. Nat. Commun. 12, 1–11 (2021).

11. Lythgoe, K. A. et al. SARS-CoV-2 within-host diversity and transmission. Science 372, eabg0821 (2021).

12. Valesano, A. L. et al. Temporal dynamics of SARS-CoV-2 mutation accumulation within and across infected hosts. PLOS Pathog. 17, e1009499 (2021).

13. Poon, L. L. M. et al. Quantifying influenza virus diversity and transmission in humans. Nat. Genet. 48, 195–200 (2016).

14. Leonard, A. S., Weissman, D. B., Greenbaum, B., Ghedin, E. & Koelle, K. Transmission Bottleneck Size Estimation from Pathogen Deep-Sequencing Data, with an Application to Human Influenza A Virus. J. Virol. 91, (2017).

15. Martin, M. A. & Koelle, K. Reanalysis of deep-sequencing data from Austria points towards a small SARS-COV-2 transmission bottleneck on the order of one to three virions. bioRxiv 2021.02.22.432096 (2021) doi:10.1101/2021.02.22.432096.

16. Varble, A. et al. Influenza A virus transmission bottlenecks are defined by infection route and recipient host. Cell Host Microbe 16, 691–700 (2014).

17. Larremore, D. B. et al. Test sensitivity is secondary to frequency and turnaround time for COVID-19 screening. Sci. Adv. 7, eabd5393 (2021).

18. Liu, T. et al. Risk factors associated with COVID-19 infection: a retrospective cohort study based on contacts tracing. Emerg. Microbes Infect. 9, 1546–1553 (2020).

19. Chen, S., Zhou, Y., Chen, Y. & Gu, J. fastp: an ultra-fast all-in-one FASTQ preprocessor. Bioinformatics 34, i884–i890 (2018).

20. Martin, M. Cutadapt removes adapter sequences from high-throughput sequencing reads. EMBnet.journal 17, 10–12 (2011).

21. Itokawa, K., Sekizuka, T., Hashino, M., Tanaka, R. & Kuroda, M. Disentangling primer interactions improves SARS-CoV-2 genome sequencing by multiplex tiling PCR. PLOS ONE 15, e0239403 (2020).

22. Li, H. & Durbin, R. Fast and accurate short read alignment with Burrows-Wheeler transform. Bioinforma. Oxf. Engl. 25, 1754–1760 (2009).

23. Grubaugh, N. D. et al. An amplicon-based sequencing framework for accurately measuring intrahost virus diversity using PrimalSeq and iVar. Genome Biol. 20, 8 (2019).

24. Hadfield, J. et al. Nextstrain: real-time tracking of pathogen evolution. Bioinformatics 34, 4121–4123 (2018).

25. Guindon, S. et al. New algorithms and methods to estimate maximum-likelihood phylogenies: assessing the performance of PhyML 3.0. Syst. Biol. 59, 307–321 (2010).

26. Yang, Z. Maximum likelihood phylogenetic estimation from DNA sequences with variable rates over sites: approximate methods. J. Mol. Evol. 39, 306–314 (1994).

27. Yu, G., Smith, D. K., Zhu, H., Guan, Y. & Lam, T. T.-Y. ggtree: an r package for visualization and annotation of phylogenetic trees with their covariates and other associated data. Methods Ecol. Evol. 8, 28–36 (2017).

