## Supplemental Table S1, Supplemental Figure S1 for "Viral infection and transmission in a large, well-traced outbreak caused by the SARS-CoV-2 Delta variant"

Table S1 iSNV pair transmission

| Transmission_pairs | iSNVs (donor) | iSNVs (recipient) | Transmitted iSNVs* |
|---|---|---|---|
| pair 1 | 3 | 2 | 1 |
| pair 2 | 3 | 2 | 1 |
| pair 3 | 3 | 1 | 1 |
| pair 4 | 19 | 3 | 0 |
| pair 5 | 1 | 4 | 0 |
| pair 6 | 1 | 49 | 0 |
| pair 7 | 1 | 0 | 0 |
| pair 8 | 1 | 44 | 0 |
| pair 9 | 1 | 44 | 0 |
| pair 10 | 1 | 2 | 0 |
| pair 11 | 21 | 2 | 0 |
| pair 12 | 3 | 5 | 0 |
| pair 13 | 3 | 0 | 0 |
| pair 14 | 21 | 0 | 0 |
| pair 15 | 21 | 5 | 0 |
| pair 16 | 21 | 1 | 0 |
| pair 17 | 21 | 19 | 0 |
| pair 18 | 21 | 3 | 0 |
| pair 19 | 21 | 6 | 1 |
| pair 20 | 21 | 25 | 0 |
| pair 21 | 3 | 0 | 0 |
| pair 22 | 3 | 5 | 0 |
| pair 23 | 3 | 1 | 0 |
| pair 24 | 3 | 19 | 0 |
| pair 25 | 3 | 3 | 0 |
| pair 26 | 3 | 6 | 0 |
| pair 27 | 3 | 25 | 0 |
| pair 28 | 1 | 1 | 0 |
| pair 29 | 1 | 0 | 0 |
| pair 30 | 1 | 2 | 0 |
| pair 31 | 1 | 5 | 0 |
| pair 32 | 1 | 1 | 0 |
| pair 33 | 1 | 1 | 0 |
| pair 34 | 1 | 0 | 0 |
| pair 35 | 1 | 3 | 0 |
| pair 36 | 1 | 2 | 0 |
| pair 37 | 1 | 0 | 0 |
| pair 38 | 1 | 0 | 0 |
| pair 39 | 1 | 2 | 0 |
| pair 40 | 1 | 1 | 0 |
| pair 41 | 1 | 2 | 0 |
| pair 42 | 1 | 0 | 0 |
| pair 43 | 1 | 2 | 0 |
| pair 44 | 44 | 2 | 0 |
| pair 45 | 6 | 3 | 0 |
| pair 46 | 6 | 3 | 0 |
| pair 47 | 6 | 8 | 0 |
| pair 48 | 6 | 1 | 0 |
| pair 49 | 6 | 1 | 0 |
| pair 50 | 6 | 4 | 0 |
| pair 51 | 25 | 1 | 0 |
| pair 52 | 25 | 6 | 0 |
| pair 53 | 25 | 2 | 2 |
| pair 54 | 25 | 1 | 1 |
| pair 55 | 25 | 0 | 0 |
| pair 56 | 25 | 15 | 0 |
| pair 57 | 25 | 2 | 0 |
| pair 58 | 25 | 1 | 0 |
| pair 59 | 25 | 2 | 0 |
| pair 60 | 5 | 1 | 0 |
| pair 61 | 1 | 3 | 1 |
| pair 62 | 1 | 4 | 0 |
| pair 63 | 1 | 4 | 0 |
| pair 64 | 2 | 1 | 0 |
| pair 65 | 2 | 3 | 0 |
| pair 66 | 2 | 0 | 0 |
| pair 67 | 1 | 2 | 0 |
| pair 68 | 1 | 1 | 0 |
| pair 69 | 1 | 2 | 0 |
| pair 70 | 1 | 3 | 0 |
| pair 71 | 4 | 3 | 0 |
| pair 72 | 1 | 3 | 0 |
| pair 73 | 4 | 0 | 0 |
| pair 74 | 4 | 4 | 0 |

* Donor iSNVs could be found in recipient.

Figure S1

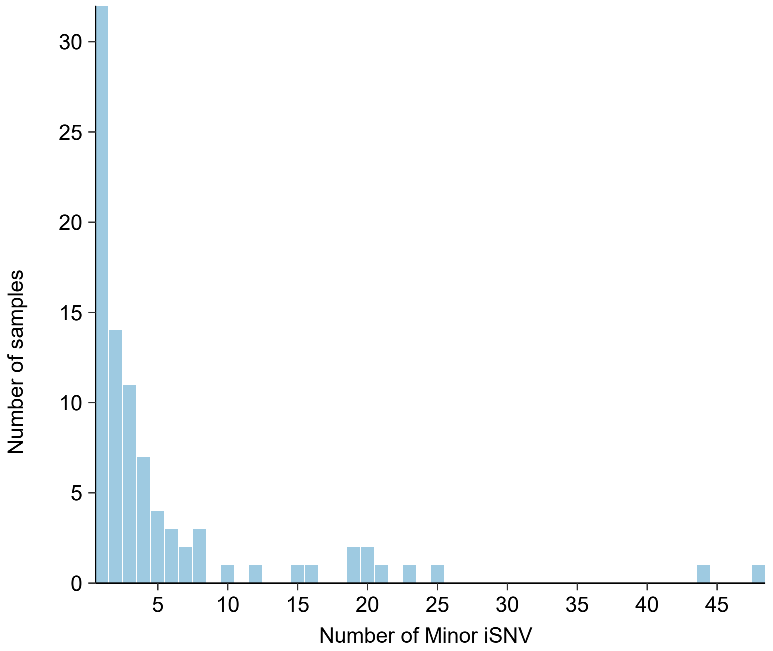


Supplemental figure S1.
Histogram of the number of samples (y-axis) by the number of minor iSNV per sample (x-axis), n = 89.
